# Prevalence, Genotype Distribution, and Determinants of High-Risk Carcinogenic Human Papillomavirus Infection Among Female Sex Workers in Kilimanjaro Region: A Community-Based Cross-Sectional Study

**DOI:** 10.1101/2025.05.01.25326808

**Authors:** Gumbo D. Silas, Federica Inturrisi, Innocent H. Uggh, Patricia Swai, Karen Yeates, Nicola West, Bariki Mchome, Alma R. Nzunda, Gaudensia Olomi, Prisca Marandu, Leah Mmari, Happiness Kilamwai, Blandina T. Mmbaga, Alex Mremi

## Abstract

**Background:** High-risk carcinogenic human papillomavirus (hrHPV) is the leading cause of cervical cancer globally. Female sex workers (FSWs) are particularly vulnerable due to high-risk sexual behaviors and low cervical cancer screening uptake. Despite their elevated risk, the prevalence, genotype distribution, and determinants of hrHPV infection among this population in Tanzania remain unknown. Hence this study.

**Methods:** A community-based cross-sectional study was conducted in Kilimanjaro region between June and July 2024, among 309 FSWs aged 25–49 years with no history of precancerous lesions or total hysterectomy. The Respondent-Driven Sampling (RDS) technique was used to recruit this hard-to-reach population. Self-collected vaginal samples were tested using the ScreenFire HPV assay, detecting 13 genotypes grouped into four risk channels. Data were analysed using SPSS Version 27.0, and a modified Poisson regression with robust error variance was applied to estimate associations between predictor variables and hrHPV infection. Statistical significance was set at p < 0.05.

**Results:** Among the 309 FSWs, the mean age was 36.1 years (SD = 5.243), with nearly half (48.5%) aged 25–34 years. The overall hrHPV prevalence was 57.6%, with higher rates among FSWs aged 40–44 years (58.5%), HIV-positive individuals (90.0%), tobacco smokers (76.8%), and those with prior cervical cancer screening (76.5%). Channel-specific hrHPV prevalence was highest for HPV31/33/35/52/58 (23.3%), followed by HPV39/51/56/59/68 (13.6%), HPV16 (9.7%), and HPV18/45 (5.5%). Mixed infections accounted for 5.5%. Religion (APR: 1.45, 95% CI: 1.13–1.61), occupation (APR: 1.28, 95% CI: 1.15–1.45), HIV status (APR: 1.27, 95% CI: 1.12–1.43), tobacco smoking (APR: 1.15, 95% CI: 1.04–1.28), and cervical cancer screening history (APR: 0.83, 95% CI: 0.73–0.94) were significantly associated with hrHPV infection in this population.

**Conclusion:** This study reveals a high prevalence of hrHPV among FSWs in Kilimanjaro, Tanzania, highlighting the urgent need for targeted cervical cancer prevention interventions for this population.

## BACKGROUND

Globally, cervical cancer remains a major health challenge, with an estimated 662,301 new cases and 348,874 deaths in 2022, accounting for 3.3% and 3.6% of all cancer cases and deaths, respectively[1]. Sub-Saharan Africa bears a disproportionately high burden, with 117,944 new cases and 76,140 deaths (13.9% and 13.6% of all cancer cases and deaths) in the same year [2]. East Africa experiences the highest regional burden, recording 58,145 new cases and 39,476 deaths accounting for 16.6% and 16.7% of all cancer cases and deaths respectively) [3]. Tanzania faces the highest burden of cervical cancer in East Africa, with 10,868 new cases and 6,832 deaths recorded in 2022, representing 24.2% of all cancer cases and 23% of cancer-related deaths in the country [4].

In 2020, the World Health Organization (WHO) launched strategies to accelerate the elimination of cervical cancer as a public health problem worldwide, setting three ambitious targets for 2030 [5]. These targets include vaccinating at least 90% of girls aged 9–14 years to protect them against high risk carcinogenic HPV, screening at least 70% of eligible women for cervical precancerous lesions at least twice in their lifetime using high-performance screening tests such as HPV DNA testing, and ensuring that at least 90% of women diagnosed with precancerous lesions receive treatment, while 90% of those with invasive cancer receive appropriate management. All WHO member states, including Tanzania, adopted these strategies.

Tanzania initially began by vaccinating all 14-year-old girls against high-risk HPV before expanding the program to include those aged 9 years[6]. The country continues to screen women aged 30–49 years for precancerous lesions using Visual Inspection with Acetic Acid (VIA), with priority given to all women including those living with HIV, who are screened starting at 25 years of age. Cryotherapy is the most common treatment for women diagnosed with precancerous lesions, while chemotherapy, surgery, and radiation therapy are the primary treatment options for those with cervical cancer disease. Ocean Road Cancer Institute serves as the only centralized national cancer treatment centre, particularly for radiotherapy.

HPV is responsible for approximately 90% of all cervical cancer cases among women, with high-risk carcinogenic genotypes contributing to over 70% of the risk of developing the disease[7]. Although all women are susceptible to HPV infection, FSWs are at significantly higher risk due to frequent exposure through multiple sexual partners, inconsistent condom use, and low cervical cancer screening uptake. This is evidenced by several studies including the systematic reviews which estimated a pooled global prevalence of HPV among this population at 42.6%, with an even higher at 79.8% in Sub-Saharan Africa, compared to the general population of women[8] [9]. This highlights the disproportionate burden faced by this marginalized population.

Despite the government’s efforts to prevent high-risk HPV infection among women, a key factor in cervical cancer, its prevalence remains unknown among FSWs, who play a significant role in transmitting the virus to the general population of women through their multiple sexual partners.

Additionally, this population is often overlooked or underserved in healthcare services, creating a gap in targeted public health interventions aimed at reducing cervical cancer incidence by focusing on high-risk groups. This gap directly impacts the United Nations’ Sustainable Development Goal (**SDG 3**), particularly **Target 3.4**, which seeks to reduce premature mortality from non-communicable diseases including cervical cancer by one-third by 2030 [5]. Additionally, the SDG’s central principle of **Leave No One Behind** also underscores the need to include marginalized populations such as FSWs in efforts towards cervical cancer elimination[10].

This study aimed to investigate the prevalence, genotype distribution, and determinants of high-risk carcinogenic human papillomavirus infection among FSWs in Kilimanjaro region, Tanzania. Therefore, to ensure equal access to cervical cancer prevention for all regardless of their social status, the findings from this study are critical for guiding the development of targeted interventions for cervical cancer prevention tailored to marginalized populations like FSWs.

Our study was nested within the NECST PAVE Tanzania study, which is part of the larger PAVE consortium aimed to evaluate the effectiveness of HPV-AVE screening within the context of the NECST initiative, which focuses on advancing cervical cancer prevention strategies. By embedding our study within this broader initiative, we leveraged the resources and protocols developed by the PAVE consortium to assess high-risk HPV prevalence and associated factors among FSWs in Kilimanjaro region.

## CONCEPTUAL FRAMEWORK

### METHODS

#### Study Design and Setting

This was a community-based cross-sectional study conducted in Kilimanjaro region, between June and July 2024. The region is located in northern Tanzania, sharing a border with Kenya to the north and East, Tanga region to the South, Manyara region to the Southwest, and Arusha region to the west. According to Tanzania’s National Population and Housing Census of 2022, the region has a total population of 1,861,934, consisting of 907,636 males and 954,298 females. Among these, 272,776 women are aged 25– 49 years, with 197,317 residing in rural areas and 75,458 in urban areas[11].The region is administratively divided into seven councils: Moshi District, Moshi Municipality, Hai District, Siha District, Rombo District, Mwanga District, and Same District. The predominant ethnic groups in this region are the Chagga and Pare who primarily engage in agricultural and trading activities. The region is also home to Mount Kilimanjaro, the highest mountain in Africa, which attracts both domestic and international tourists.

#### Study Population, Sample Size, and Sampling Technique

A total of 309 FSWs aged 25-49 years with no prior history of pre-cancerous lesions, nor total hysterectomy (as they would not have a cervix for sampling), and who had been engaging in commercial sex for at least 6 months before the commencement of this study, were recruited from hotspots located in Moshi District and Moshi Municipal Councils in Kilimanjaro region. We used a Respondent-Driven Sampling, a peer-referral technique commonly used to recruit the hard-to-reach populations like FSWs whereby normal sampling techniques are usually not effective. RDS builds recruitment chains from initial participants called “seeds”, and applies statistical adjustments to reduce biases, enhance confidentiality, and allow accurate estimation of population characteristics[13].

The sample size (309) was estimated using Cochran’s Sample Size Formula, considering a 95% confidence level (Z-score), an estimated prevalence of high-risk HPV (HrHPV) of 23.6% based on a previous study conducted among the same population in Kenya, and a precision level (ꜫ). To account for potential non-response, the calculated sample size was increased by **10%** [14].

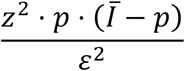

#### Data collection procedure

An interviewer-administered questionnaire was used to collect social demographic information and sexual behaviors of study participants, with the National Key and Vulnerable Population (KVP) screening tool used to confirm the key inclusion criteria of being engaging in commercial sex for at least 6 months prior to the commencement of this study. HPV samples were self-collected and transported to the pathology laboratory at Kilimanjaro Christian Medical Centre tertiary zonal referral hospital. The samples were tested using ScreenFire HPV RS assay, a diagnostic test designed for the detection and identification of human papillomavirus (HPV) DNA[15]. It utilizes isothermal DNA amplification and is the first HPV genotyping assay specifically developed to deliver rapid and cost-effective results. The assay organizes outcomes into four channels based on the risk of causing cervical cancer. It targets HPV16 (channel1), HPV18/45(channel 2), HPV31/33/35/52/58(channel 3), and HPV39/51/56/59/68 (channel 4)[16].

Participants who tested positive for high-risk carcinogenic HPV were notified confidentially via their mobile phones and linked to triage screening under the framework of the NECST PAVE Tanzania study. As part of this approach, participants underwent further diagnostic procedures, including cervical examination through visual inspection with acetic acid (VIA) and biopsy when indicated. Management and treatment were provided based on the results, following local standard guidelines aligned with the NECST study protocols. Trained peer educators played a critical role in facilitating participant navigation and provided support during follow-up appointments.

#### Data analysis

Data analysis was carried out using SPSS Version 27.0. In descriptive statistics, categorical variables were summarized into frequencies and percentages, and continuous variables in means and standard deviations. In inferential analysis, a Chi-square test was performed to assess the associations between categorical variables and the prevalence of high-risk Carcinogenic HPV, followed by a modified Poisson regression model with robust error variance, which was applied to determine the strength of the observed associations. Weighting adjustments were performed to minimize the potential biases resulting from RDS. All variables with P-values ≤0.2 in bivariate analysis were included in multivariate analysis and only those with p-values below 0.05 in multivariate analysis were considered statistically significant.

## RESULTS

### Sociodemographic and behavioural Characteristics of Study Participants

This study enrolled 309 FSWs with an average age of 36.11 years and a standard deviation of 5.243. Nearly half of the participants were aged between 25-34 years150 (48.5%), and the majority resided in urban areas, 194 (62.8%). The vast majority were involved in peasantry alongside commercial sex work 297 (96.1%). Most participants identified as Christians 282 (91.3%), and primary was the highest level of education attained by the majority 167 (54.0%). Nearly half of the participants were separated or divorced151(48.9%), with most having three or more children 207 (67.0%). The majority were HIV-negative, 279 (90.3%), and most initiated sexual activity between the ages of 15-19 years 261(84.5%). The use of hormonal Contraceptives was reported by 238(77.0%) of the participants, while 56 (18.1%) were tobacco smokers. Most of the participants 162(52.5%) started engaging in commercial sex between the age of 15-19 years with only 17(5.5%) having a history of cervical cancer screening (Table 1).

**Table 1.** Sociodemographic and behavioural characteristics of Study Participants (N=309)

| Variable | Frequency | Percentage | P-value |
| --- | --- | --- | --- |
| **Age (Years)** | Mean±SD 36.11±5.243 |  | 0.946 |
| 25-34 | 150 | **48.5** |  |
| 35-39 | 83 | 26.9 |  |
| 40-44 | 53 | 17.2 |  |
| 45-49 | 23 | 7.4 |  |
| **Residence** |  |  | 0.873 |
| Rural | 115 | 37.2 |  |
| Urban | 194 | **62.8** |  |
| **Occupation** |  |  | 0.678 |
| FSW+Business | 1 | 0.3 |  |
| FSW+Peasants | 297 | **96.1** |  |
| FSW+Housewife | 11 | 3.6 |  |
| **Religion** |  |  | 0.500 |
| Others | 1 | 0.3 |  |
| Christians | 282 | **91.3** |  |
| Muslims | 26 | 8.4 |  |
| **Education** |  |  | 0.529 |
| Never went to school | 13 | 4.0 |  |
| Primary | 167 | **54.0** |  |
| Secondary | 120 | 38.8 |  |
| College/University | 9 | 2.9 |  |
| **Marital Status** |  |  | 0.120 |
| Single | 102 | 33.0 |  |
| Married/Cohabiting | 56 | 18.1 |  |
| Separated/Divorced | 151 | **48.9** |  |
| **Parity** |  |  | 0.133 |
| Nulliparous | 25 | 8.1 |  |
| Para 1-2 | 77 | 24.9 |  |
| Para 3+ | 207 | **67.0** |  |
| **HIV status** |  |  | <0.001 |
| Negative | 279 | **90.3** |  |
| Positive | 30 | 9.7 |  |
| **Age sexual debut** |  |  | 0.912 |
| 15-19 Years | 48 | **84.5** |  |
| 20+ Years | 261 | 15.5 |  |
| **Use contraceptives** |  |  | 0.428 |
| Yes | 238 | **77.0** |  |
| No | 71 | 23.0 |  |
| **Tobacco Smoking** |  |  | 0.003 |
| Yes | 56 | 18.1 |  |
| No | 253 | **81.9** |  |
| **Age started sex work** |  |  | 0.130 |
| <15 | 6 | 1.9 |  |
| 15-19 | 141 | **52.5** |  |
| 20-24 | 162 | 45.6 |  |
| **History of Screening** |  |  | 0.003 |
| No | 292 | **94.5** |  |
| Yes | 17 | 5.5 |  |

#### Prevalence of high-risk carcinogenic HPV among FSWs in Kilimanjaro region

The overall prevalence of any high-risk carcinogenic HPV among this population was 178(57.6%), with no significant differences observed across age groups, residential areas, sexual debut and condom use. FSWs involved in peasantry exhibited the highest prevalence of 171(57.6%) compared to other occupations, while those with secondary education had a slightly higher prevalence 75(62.5%) compared to other education levels. The prevalence was also high among HIV positive FSWs 23(90.0%) compared to their HIV-negative counterparts 155(54.1%). Tobacco smokers also exhibited a markedly higher prevalence of 135(76.8%) compared to non-smokers 43(53.4%). Furthermore, those who had previously undergone cervical cancer screening demonstrated a higher prevalence of 13(76.5%) compared to those who had never undergone screening 118(40.4) (Table 2).

**Table 2:** Prevalence of any high-risk Carcinogenic Human papillomavirus among FSWs (N=309)

| Variable | HPV Status n(%) |  | P-value |
| --- | --- | --- | --- |
|  | Positive | Negative |  |
| **Age (Years)** |  |  |  |
| 25-34 | 87(58.0) | 63(42) |  |
| 35-39 | 47(56.6) | 36(43.4) |  |
| 40-44 | 31(58.5) | 22(41.5) |  |
| 45-49 | 13(56.5) | 10(43.5) |  |
| **Residence** |  |  |  |
| Rural | 67(58.3) | 48(41.7) |  |
| Urban | 111(57.2) | 83(42.8) |  |
| **Religion** |  |  |  |
| Others | 1(100) |  |  |
| Christians | 164(58.2) | 118(41.8) | <0.001 |
| Muslims | 13(50.0) | 13(50.0) | <0.001 |
| **Occupation** |  |  |  |
| FSW+Business | 1(100.0) | 0(0.00) |  |
| FSW+Peasants | 171(57.6) | 126(42.4) | <0.001 |
| FSW+Housewife | 6(54.5) | 5(45.5) | 0.018 |
| **Education** |  |  |  |
| Never | 7(53.8) | 6(46.2) |  |
| Primary | 91(54.5) | 76(45.5) |  |
| Secondary | 75(62.5) | 45(37.5) |  |
| College/University | 5(55.6) | 4(44.4) |  |
| **Marital Status** |  |  |  |
| Single | 67(65.7) | 35(34.3) |  |
| Married/Cohabiting | 25(44.6) | 31(55.4) |  |
| Separated/Divorced | 86(57.0) | 65(43.0) |  |
| **Parity** |  |  |  |
| Nulliparous | 18(72.0) | 7(28.0) |  |
| Para 1-2 | 46(59.7) | 31(40.3) | 0.119 |
| Para 3+ | 114(55.1) | 93(44.9) | 0.163 |
| **HIV status** |  |  |  |
| Negative | 155(54.1) | 124(45.9) |  |
| Positive | 23(90.0) | 7(10.0) | <0.001 |
| **Sexual debut** |  |  |  |
| 15-19 Years | 28(58.3) | 20(41.7) |  |
| 20+ Years | 150(57.5) | 111(42.5) |  |
| **Tobacco Smoking** |  |  |  |
| No | 43(53.4) | 13(23.2) |  |
| Yes | 135(76.8) | 118(46.6) | 0.006 |
| **Use condom** |  |  |  |
| Yes | 100(41.7) | 140(58.3) |  |
| No | 31(44.9) | 38(55.1) |  |
| **History Screening** |  |  |  |
| No | 118(40.4) | 174(59.6) |  |
| Yes | 13(76.5) | 4(23.5) | 0.003 |

This study also observed a channel specific prevalence which consistently increased with age in three of the channels except channel 1. For Channel 1 (HPV16), the prevalence increased from 1.3% among those aged 25–34 years to 3.6% among those aged 40–44 years, before slightly decreasing to 2.9% in the 45–49-year group. For Channel 2 (HPV18/45), the prevalence steadily increased from 0.3% in the 25–34-year group to 13.2% in the 45–49-year group. Channel 3 (HPV31/33/35/52/58) showed a steady increase, from 1.6% in the 25–34-year group to 12.6% in the 45–49-year group. Similarly, Channel 4 (HPV39/51/56/59/68) showed an increase in prevalence, from 1.0% in the 25–34-year group to 6.1% in the 45–49-year group. For Mixed Infections, also the prevalence increased from 0.3% among 25-34 years to 3.6% among those aged between 45–49-years (Figure 3).

**Figure 1:**
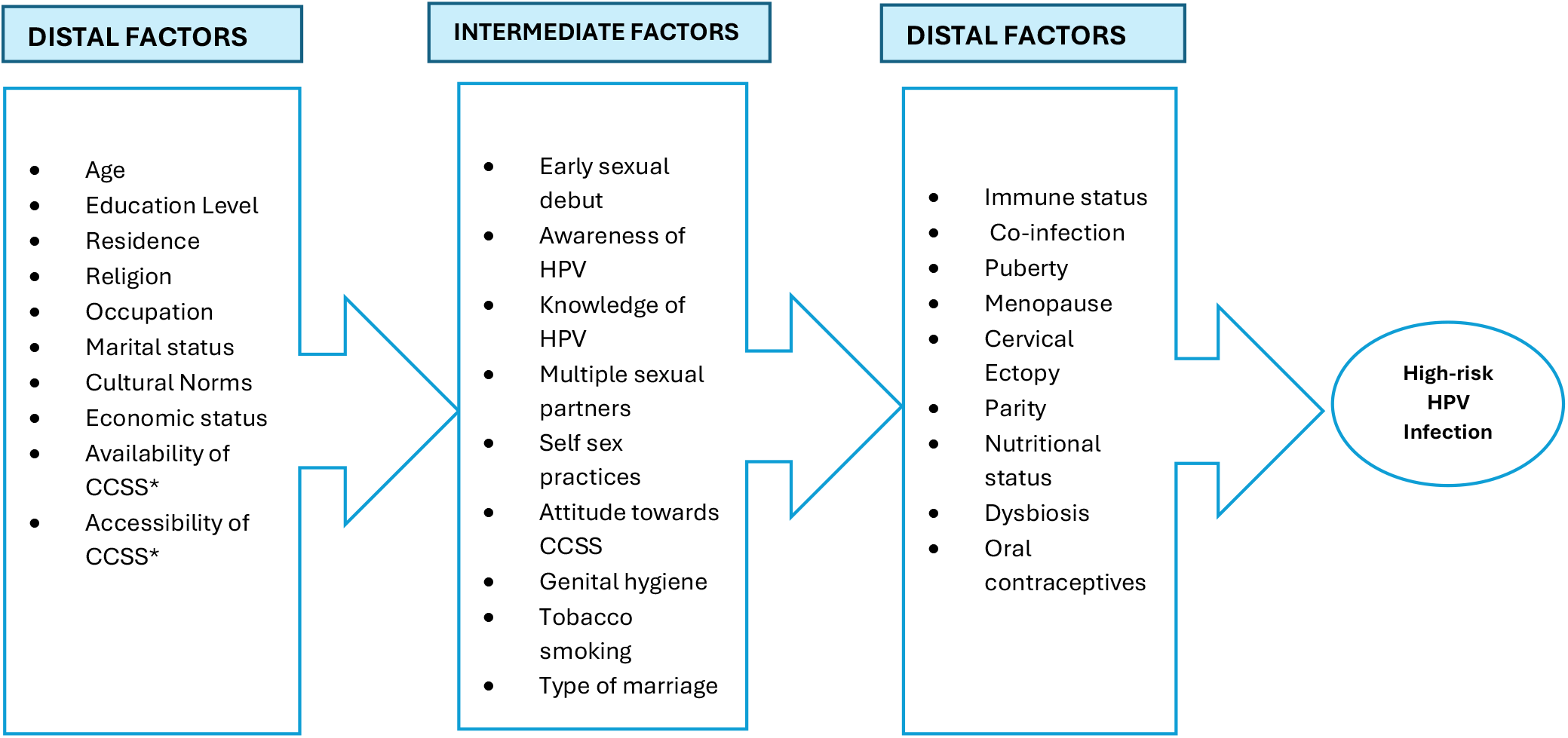
Factors influencing the rate of infection with high-risk carcinogenic HPV) among FSWs * (CCSS=Cervical cancer Screening Services)

**Figure 2:**
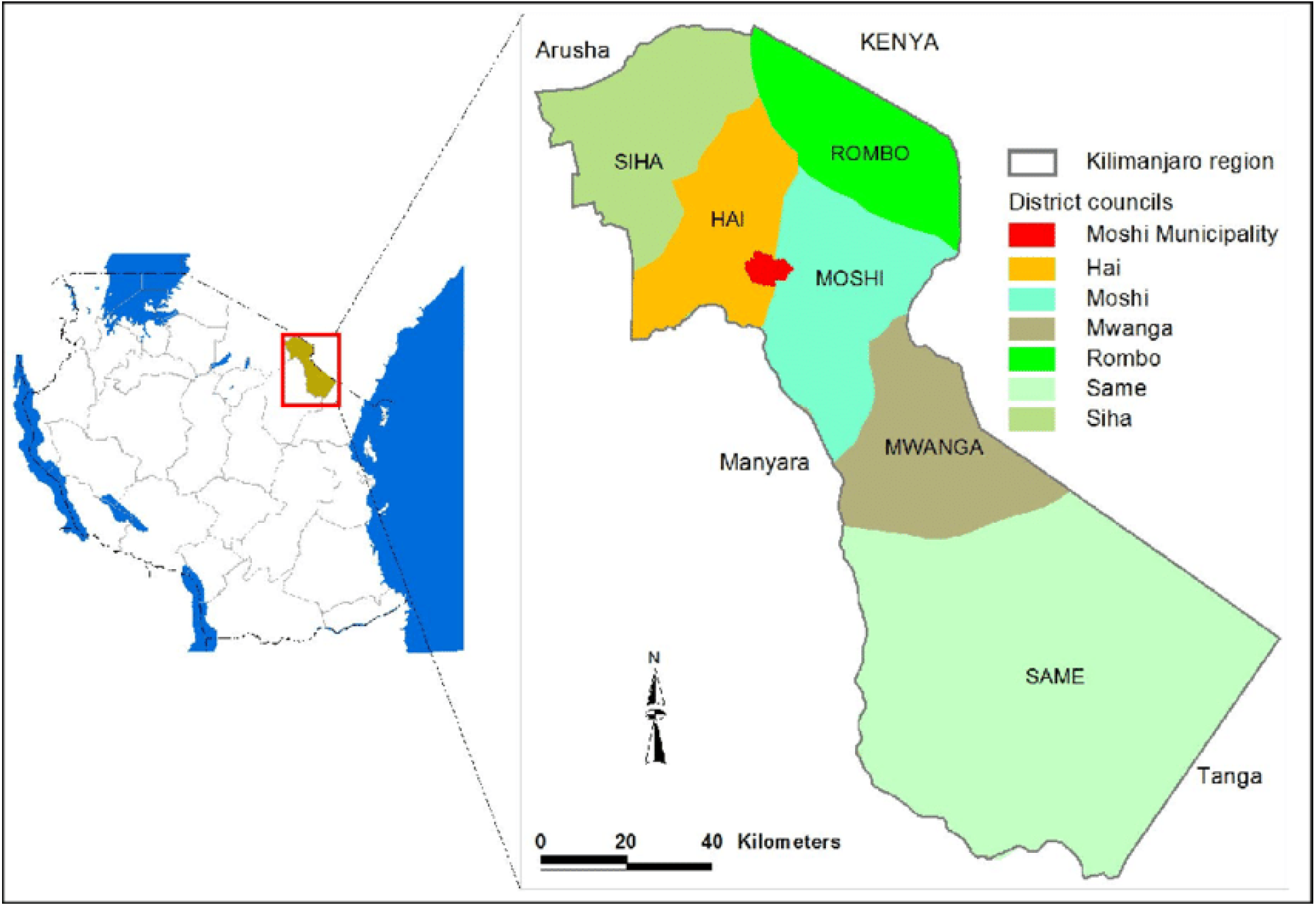
The map of Kilimanjaro region showing the borders and Councils [12]

**Figure 3:**
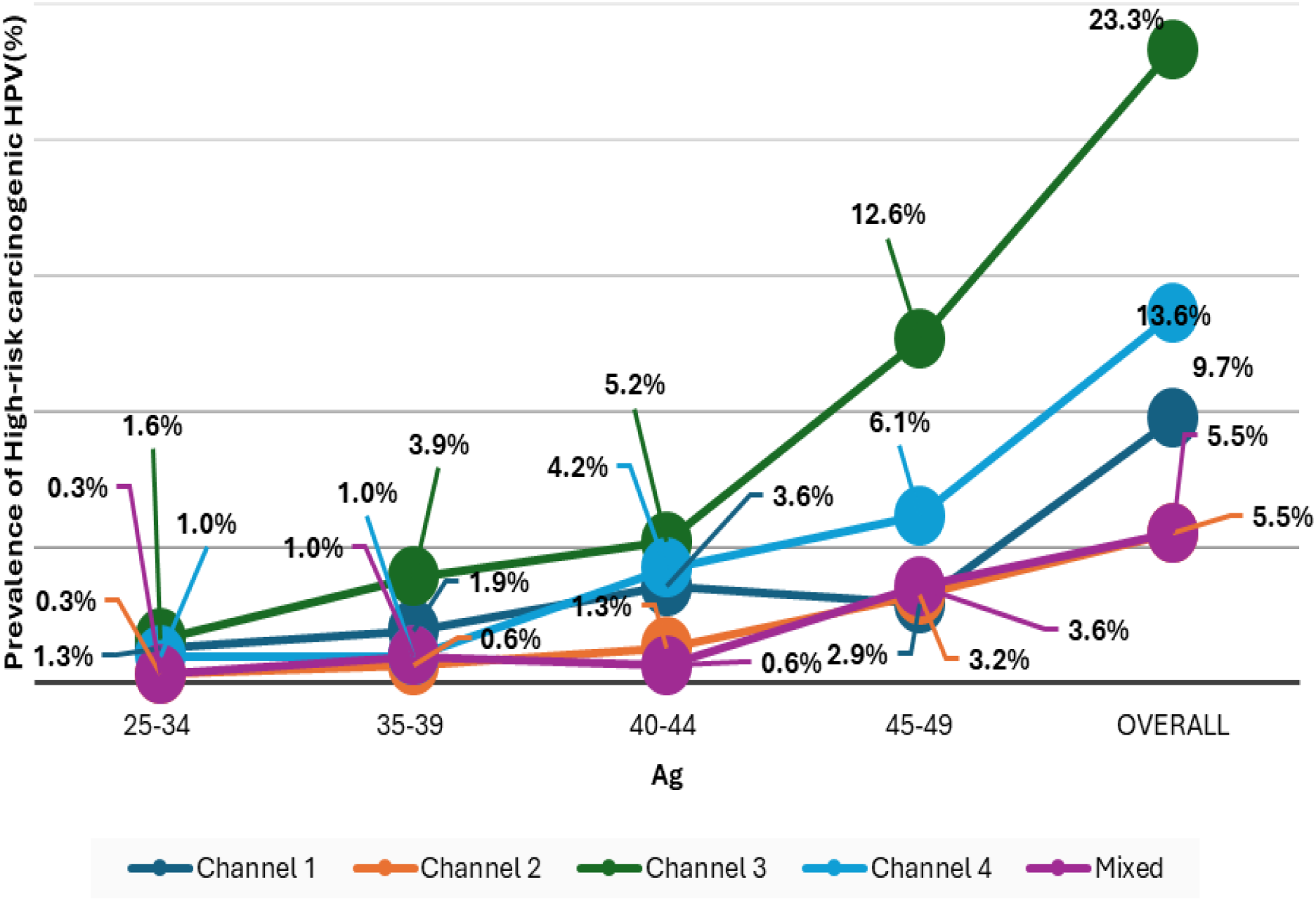
Distribution of Prevalence of specific genotypes of High-risk Carcinogenic HPV by age (N=309)

#### Distribution of channel specific High-risk Carcinogenic HPV cases among Study participants by age

This study revealed a varying distribution of high-risk carcinogenic HPV genotypes among this population, with HPV31/33/35/52/58 (Channel 3) exhibiting the highest number of cases72(23.3%), followed by 42 cases (13.6%) in HPV39/51/56/59/68 (Channel 4), 30 cases (9.7%) in HPV16 (Channel 1), while both HPV18/45 (Channel 2) and Mixed Infections, accounted for 17 cases (5.5%) each (Figure 3&4).

**Figure 4:**
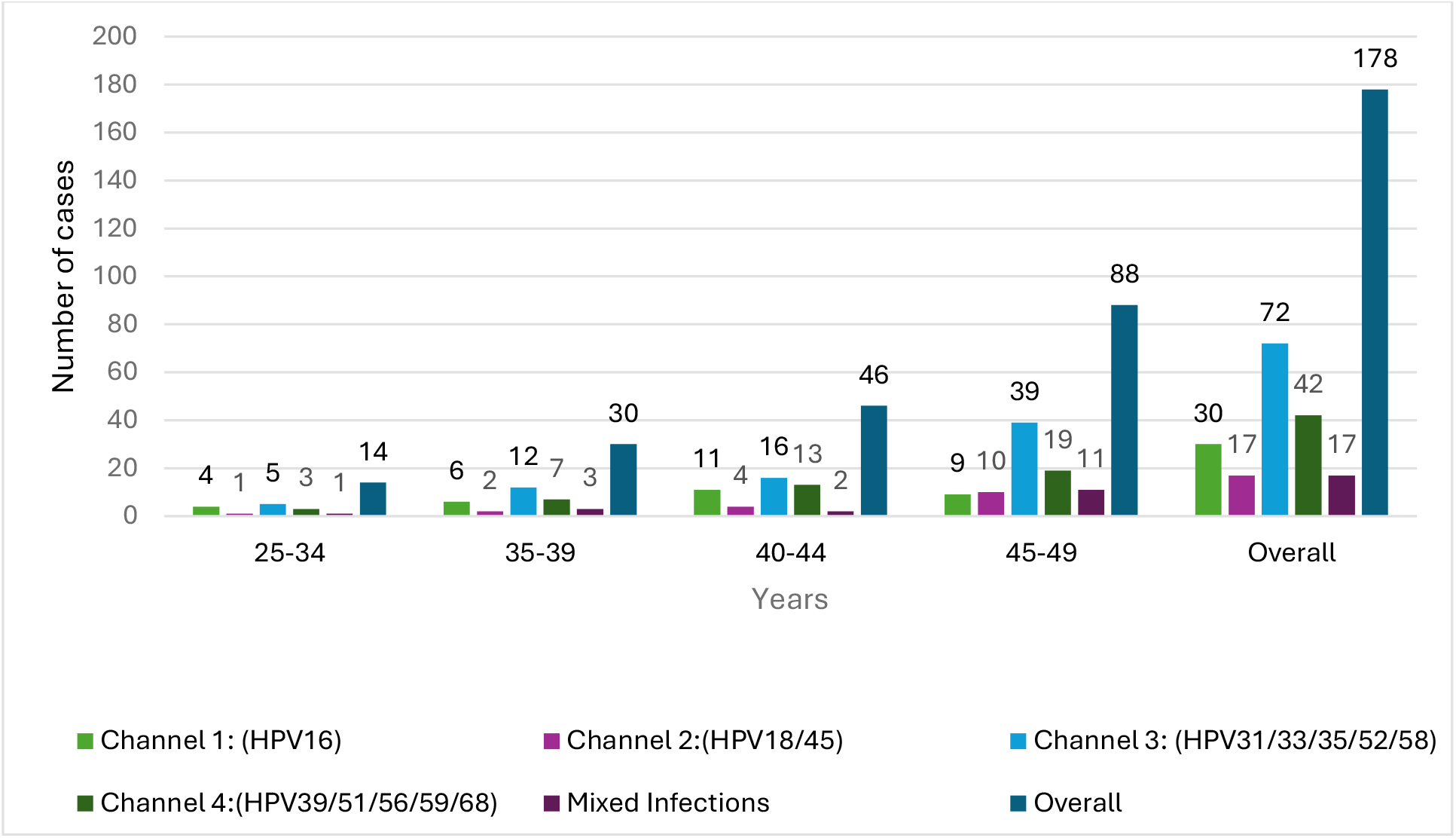
Distribution of channel specific High-risk carcinogenic HPV among FSWs by age (N=178)

### Determinants of high-risk carcinogenic HPV infections among FSWs in Kilimanjaro

In this study, religion, occupation, HIV status, tobacco smoking, and history of cervical cancer screening were significantly associated with high-risk carcinogenic HPV infection among this population.

Participants who identified as Christians (APR: 1.45, 95% CI: 1.13–1.61) and Muslims (APR: 1.15, 95% CI: 1.30–1.74) had 45% and 15% increased likelihood of being infected with high-risk carcinogenic HPV respectively, compared to those practising other religions. Regarding occupation, FSWs involved in peasantry (APR: 1.28, 95% CI: 1.15–1.45) and those who were housewives (APR: 1.26, 95% CI: 1.04–1.53) exhibited a 28% and 26% increased likelihood of being infected with high-risk-carcinogenic HPV respectively, compared to those engaged in other businesses (Table 3).

**Table 3:** Determinants of high-risk carcinogenic Human papillomavirus infection among FSWs (N=309)

| **Variable** | **Bivariate Analysis**<br>**CPR(95% CI)** | **P-value** | **Multivariate Analysis**<br>**APR (95% CI)** | **P-value** |
| --- | --- | --- | --- | --- |
| **Age (Years)** |  |  |  |  |
| 25-34 | 1 |  |  |  |
| 35-39 | 0.94(0.67-1.31) | 0.708 |  |  |
| 40-44 | 1.01(0.72-1.41) | 0.965 |  |  |
| 45-49 | 1.02(0.67-1.55) | 0.937 |  |  |
| **Residence** |  |  |  |  |
| Rural | 1 |  |  |  |
| Urban | 0.99(0.91-1.07) | 0.751 |  |  |
| **Religion** |  |  |  |  |
| Others | 1 |  | 1 |  |
| Christians | 1.52(1.44-1.61) | 0.000 | 1.45(1.13-1.61) | <0.001 |
| Muslims | 1.65(1.36-1.99) | <0.001 | 1.15(1.30-1.74) | <0.001 |
| **Occupation** |  |  |  |  |
| FSW+Business | 1 |  | 1 |  |
| FSW+Peasants | 1.53(1.45-1.62) | 0.000 | 1.28(1.15-1.45) | <0.001 |
| FSW+Housewife | 1.58(1.17-2.11) | 0.002 | 1.26(1.04-1.53) | 0.018 |
| **Education** |  |  |  |  |
| Never | 1 |  |  |  |
| Primary | 0.01(0.76-1.35) | 0.937 |  |  |
| Secondary | 1.005(0.79-1.27) | 0.966 |  |  |
| College/University | 1.01(0.76-1.35) | 0.937 |  |  |
| **Marital Status** |  |  |  |  |
| Single | 1 |  |  |  |
| Married/Cohabiting | 1.07(0.97-1.03) | 0.170 |  |  |
| Separated/Divorced | 0.95(0.87-1.19) | 0.215 |  |  |
| **Parity** |  |  |  |  |
| Nulliparous | 1 |  | 1 |  |
| Para 1-2 | 1.14(0.98-1.33) | 0.096 | 1.14(0.97-1.34) | 0.119 |
| Para 3+ | 1.17(1.01-1.35) | 0.032 | 1.12(0.96-1.29) | 0.163 |
| **HIV status** |  |  |  |  |
| Negative | 1 |  | 1 |  |
| Positive | 1.94(1.91-1.99) | 0.000 | 1.27(1.12-1.43) | <0.001 |
| **Sexual debut** |  |  |  |  |
| 15-19 Years | 1 |  |  |  |
| 20+ Years | 0.99(0.89-1.12) | 0.912 |  |  |
| **Tobacco Smoking** |  |  |  |  |
| No | 1 |  | 1 |  |
| Yes | 1.18(1.07-1.29) | 0.001 | 1.15(1.04-1.28) | 0.006 |
| **Use condom** |  |  |  |  |
| Yes | 1 |  |  |  |
| No | 1.02(0.93-1.12) | 0.628 |  |  |
| **History Screening** |  |  |  |  |
| No | 1 |  | 1 |  |
| Yes | 1.26(1.11-1.42) | <0.001 | 0.83(0.73-0.94) | 0.003 |

Additionally, HIV-positive participants (APR: 1.27, 95% CI: 1.12–1.43) and tobacco smokers (APR: 1.15, 95% CI: 1.04–1.28) had 27% and 15% increased likelihood of being infected with high-risk carcinogenic HPV compared to HIV-negative participants and non-smokers, respectively. Furthermore, participants with a history of cervical cancer screening had a 17% reduced likelihood of being infected with high-risk carcinogenic HPV (APR: 0.83, 95% CI: 0.73–0.94) compared to those who had never been screened (Table 3).

## DISCUSSION

This study reveals a high prevalence (57.6%) of high-risk carcinogenic HPV among FSWs in Kilimanjaro Region, with notable genotype distribution and associations with religion, occupation, HIV status, tobacco smoking, and history of cervical cancer screening. The observed high prevalence can be attributed to increased exposure to sexually transmitted infections through engagement in unprotected sexual practices with multiple partners, coupled with low awareness and knowledge about HPV as the necessary cause of cervical cancer.

Our findings are consistent with several studies conducted among the same population in various countries. For instance, a systematic review and meta-analysis involving 21,402 FSWs from 62 studies across 33 countries worldwide reported a pooled prevalence of 42.6%[8]. Similarly, a study in Cameroon reported a prevalence of 62.1% [17], while another in Nigeria revealed an even higher prevalence of 84%[18]. Furthermore, a cross-sectional study conducted among 665 FSWs in West Africa, including Benin and Mali, found prevalences of 95.5% and 81.4%, respectively [19]. Additionally, a systematic review and meta-analysis of 107 studies from 45 countries reported a pooled prevalence of 39.5%, which was noted to be approximately four times higher than that of the general population of women[20].

In contrast, lower prevalence rates were reported by two cross-sectional studies conducted in Vietnam and Greater Accra, Ghana, involving 669 and 109 FSWs, which revealed prevalences of 17.6% and 26%, respectively[18,19]. Differences in sample size, geographical location, and demographic characteristics of the study populations may explain these discrepancies. Additionally, variations in HPV screening methods, sexual practices, and differing levels of access to healthcare services could contribute to the observed differences. For example, regions with higher HIV prevalence among FSWs may also exhibit higher HPV prevalence due to the synergistic relationship between HIV and HPV infections.

This regional observation can be explained by similar reasons of increased exposure to multiple sexual partners, which significantly raises the risk of acquiring and sustaining high risk carcinogenic HPV infections among this population [23]. Given the frequent sexual activity inherent in commercial sex, FSWs are continuously exposed to different HPV strains, particularly in environments with limited condom use. Another possible explanation could be limited access to preventive services, as many FSWs face structural barriers such as stigma, discrimination, and healthcare exclusion, limiting their engagement in preventive care[19,20].

This study revealed a significant pattern in the distribution of high-risk carcinogenic HPV genotypes grouped into four channels. Notable variations exist, with certain genotypes exhibiting higher cases and prevalence across different age groups. Channel 3 genotypes showed the highest number of cases and overall prevalence (23.3%), followed by Channel 4 (13.6%) and HPV16 (Channel 1, 9.7%), while HPV18/45 (Channel 2, 5.5%) and mixed infections (5.5%) displayed relatively lower prevalence.

The increasing prevalence of these genotypes with age suggests that older FSWs are more likely to harbours high-risk HPV infections. This trend could be attributed to cumulative exposure over time and persistence of infections acquired earlier in life. Additionally, compromised immune systems, such as those observed in HIV-infected individuals, may reduce the body’s ability to clear the virus, leading to persistent infections more likely to progress to high-grade lesions and, eventually, cervical cancer if not adequately managed.

HPV16 exhibited an increase in cases with age, peaking among those aged 40–44 years, followed by a slight decline in the 45–49-year group. This decline could be due to natural clearance of the virus, progression to advanced disease stages where detection is more challenging, or a reduced likelihood of new infections in older age groups. Mixed infections, though representing a smaller proportion of cases, increased with age, with the highest number of cases observed in the oldest age group. This pattern may be associated with prolonged co-infections resulting from weakened immune responses and ongoing high-risk behaviours.

The prevalence of specific HPV genotypes among FSWs observed in this study aligns with several studies conducted in different regions, although variations exist. A systematic review and meta-analysis involving 107 studies from 45 countries worldwide, reported HPV16 (9%), HPV52 (8.3%), and HPV58 (6.2%) as common genotypes among FSWs, consistent with our observation of HPV52 and HPV58 being predominant in Channel 3 [26]. The relatively high prevalence of HPV52 reported in both studies further supports its significant burden among FSWs.

Similar findings were reported in a cross-section study from Kenya, where HPV52 (10.1%) was the most prevalent genotype within Channel 3. Additionally, their identification of HPV35 (2.3%) and HPV51 (2.3%) within Channels 3 and 4, respectively, aligns with our findings of moderate prevalence within these channels[14]. Moreover, a cross-sectional study involving 100 FSWs in Ghana, similarly identified HPV16 (8%) and HPV35 (5%) within Channels 1 and 3, respectively. Although the prevalence of HPV35 was slightly lower compared to our findings, the detection of mixed infections (5%) was similar to our study’s reported mixed infection rate of 5.5%[27].

Contrastingly, some studies reported higher prevalence rates of specific genotypes compared to our findings. For instance, a study conducted in West Africa (Benin and Mali) involving 659 FSWs reported significantly higher prevalence rates of HPV16 (36.6%) and HPV52 (28.8%) which are under Channels 1 and 3, respectively. Additionally, HPV58 (37.5%) was notably higher compared to our findings of 23.3% within Channel 3[19]. Similarly, a cross-sectional study involving 436 FSWs in Senegal, reported exceptionally high prevalence rates of HPV52 (32.6%), HPV35 (16.1%), HPV31 (12.4%), and HPV33 (12.8%), all classified under Channel 3[9]. Furthermore, a similar cross-sectional study from Vietnam involving 699 FSWs reported considerably lower prevalence rates of HPV16/18 (4%), HPV52 (7%), and HPV58 (6%)[21]. These disparities could be attributed to differences in study population characteristics, sample size, regional epidemiology, sample collection techniques and diagnostic assays used.

Regarding determinants, religion, occupation, HIV status, tobacco smoking, and history of cervical cancer screening were significantly associated with high-risk carcinogenic HPV infection among this population. Religion was significantly associated with high-risk carcinogenic HPV infection among this population, with those practising Christianity and Islam exhibiting a 45% and 15% increased risk of infection compared to FSWs practising other religions respectively. This can be explained by differences in cultural norms associated with different religious groups which may influence an individual’s health-seeking behaviour. FSWs practising Christianity and Islam may have less access to HPV preventive services due to stigma or restrictive beliefs within their communities; for instance, some religious norms discourage condom use. This may increase the risk of sexually transmitted infections, including high-risk carcinogenic HPV. Our finding is indirectly supported by a cross-sectional study conducted in Saudi Arabia and Texas which reported a significant proportion of women opposing the HPV vaccine due to religious reasons. Refusal to accept HPV vaccination can indirectly influence the infection with high-risk carcinogenic HPV infection [28] [29].

Occupation combination emerged as a significant factor for high-risk carcinogenic HPV among FSWs, with those who were involved in commercial sex alongside peasantry and housewives exhibiting a 28% and 26% increased risk of high-risk carcinogenic HPV respectively. This could be attributed to several factors. Peasant FSWs may have limited access to healthcare services due to residing in rural or remote areas where HPV preventive services like screening are less accessible. For housewives, societal stigma and cultural norms may discourage seeking reproductive health services, especially when engaged in sex work secretly.

HIV status emerged as a third significant factor, with HIV-positive FSWs exhibiting a higher risk of carcinogenic HPV infection compared to their HIV-negative counterparts. This observation can be explained by the immunosuppressive nature of HIV, which impairs the ability to clear HPV infections which in turn leads to persistency. Our finding was consistent with several studies done in different regions. For instance, a cross-sectional study done among 665 FSWs in West Africa revealed a 26% increase in the risk of high risk carcinogenic HPV infection among HIV-positive FSWs in Benin and Mali[19]. Similarly, two cross-sectional studies done in Kenya, Nigeria, and Senegal and a scoping review conducted in eastern and southern Africa also revealed a significant association of HIV with High-risk Carcinogenic HPV[30,31][9][32]. This indicates that HIV infection exacerbates the likelihood of developing HPV infection as a result of immune suppression[33]. Conversely, our findings were not similar to two studies done in Zimbabwe which reported no significant association between HIV status and HPV prevalence[30,31]. This could be due to the differences in study settings, sample sizes, or the prevalence of co-infections and variations in healthcare access.

Tobacco smoking emerged as a fourth significant risk factor for high-risk carcinogenic infection among this population. This may be because smoking compromises immune function. Smoking could lead to chronic inflammation, creating a favourable environment for HPV persistence and progression to cervical cancer[36]. Our finding was consistent with the prospective cohort and a cross-sectional study done in China and Jarkata, which both revealed that exposure to tobacco smoking was associated with an increased risk of developing cervical cancer through interference with the immune response to HPV infection. [37,38].

History of cervical cancer screening was a fifth significant factor for high-risk carcinogenic HPV among FSWs in this study, with FSWs who had ever been screened for cervical cancer in their lifetime having a 17% reduced risk of being infected compared to those who had never. This may be due to regular screening which facilitates early detection and management of HPV-related abnormalities, allowing for timely interventions that may prevent persistent infections and progression to cervical cancer[39]. Our finding was consistent with findings from a cross-sectional study nested in a 5-year cohort study in Denmark, which reported that women with a history of regular cervical cancer screening were less likely to be infected with high-risk carcinogenic HPV compared to those who had never been screened[40].

## CONCLUSIONS

The prevalence of any high-risk and channel-specific carcinogenic HPV among FSWs in Kilimanjaro region was significantly high. This was coupled with a varied distribution of specific genotypes, with religion, occupation, HIV status, tobacco smoking, and history of cervical cancer screening identified as significant factors contributing to the noted high prevalence. These findings underscore the need for targeted interventions aimed at enhancing access to regular cervical cancer screening and promoting HPV vaccination among FSWs to reduce the burden of high-risk carcinogenic HPV infections and prevent HPV-related cervical cancer. Such efforts are crucial, considering the role of this high-risk population in the continued transmission of the virus to the general population through their sexual partners.

However, the variation in genotype prevalence by age remains underexplored in many studies, indicating a significant gap in the literature. Furthermore, to the best of our knowledge, no previous studies have specifically reported an increased risk of high-risk carcinogenic HPV infection among FSWs engaged in commercial sex alongside peasantry or housewife duties. Therefore, further research is warranted to confirm these findings, explore the underlying mechanisms, and assess genotype-specific prevalence by age to better understand risk profiles and inform tailored preventive interventions.

### Strengths of the study

- This is the first study focusing specifically on FSWs, a marginalized population that is often overlooked.
- It lays the foundation for future research and the development of interventions aimed at improving cervical cancer prevention among marginalised populations in Tanzania.

### Limitations of the study

- Our Study revealed a consistent increase in the prevalence of HrHPV with age across all channels. This trend lacks an evidence-based explanation. Therefore, future research should consider investigating age-related factors contributing to this pattern.

## Data Availability

All data produced in the present study are available upon reasonable request to the authors

## Abbreviations

APR: Adjusted Prevalence Ratio
CPR: Crude Prevalence Ratio
DC: District Council
DNA: Deoxyribonucleic Acid
HIV: Human Immunodeficiency Virus
HPV: Human Papillomavirus
HrHPV: High Risk Human Papillomavirus
KCMC: Kilimanjaro Christian Medical Centre
KCMUCo: Kilimanjaro Christian Medical University College
KCRI: Kilimanjaro Clinical Research Institute
KVP: Key and Vulnerable Population
MDC: Moshi District Council
MMC: Moshi Municipal Council
PAVE: HPV-Automated Visual Evaluation (AVE)
RDS: Respondent-Driven Sampling
SPSS: Statistical Package for Social Sciences
TAWREF: Tanzania Women Research Foundation
VIA: Visual Inspection with Acetic Acid
WHO: World Health Organization

## Declarations

### Ethical Approval

This study complied fully with ethical standards. Ethical clearance (No. PG02/2024) was granted by Kilimanjaro Christian Medical University College (KCMUCo), with approval letters: DC.109/228/01/K, MMC/A.40/13/VOL.V/138 and MDC/M.10/18/VOL.III/17 obtained from regional, Moshi Municipal, and Moshi District authorities respectively.

### Patient consent for publication

All participants in this study gave consent.

### Availability of Data and Materials

The datasets used and/or analysed during the current study are available from the corresponding author on reasonable request.

### Authors’ contribution

- Gumbo D. Silas (*First author*): Conceptualization, Methodology, Data Collection, Data Analysis, Writing—Original Draft, Writing—Review & Editing.
- Federica Inturrisi: Data Analysis, Statistical Support, Writing—Review & Editing.
- Innocent H. Uggh: Data Collection, Investigation, Writing—Review & Editing.
- Patricia Swai: Data Collection, Supervision, Writing—Review & Editing.
- Karen Yeates: Conceptualization, Methodology, Funding Acquisition, Writing—Review & Editing.
- Melinda Chelva: Methodology, Investigation, Writing—Review & Editing.
- Nicola West: Methodology, Writing—Review & Editing.
- Bariki Mchome: Data Collection, Writing—Review & Editing.
- Alma R. Nzunda: Data Collection, Writing—Review & Editing.
- Gaudensia Olomi: Data Collection, Writing—Review & Editing.
- Prisca Marandu: Data Collection, Writing—Review & Editing.
- Leah Mmari: Data Collection, Writing—Review & Editing.
- Happiness Kilamwai: Data Collection, Writing—Review & Editing.
- Blandina T. Mmbaga: Conceptualization, Methodology, Supervision, Funding Acquisition, Writing—Review & Editing.
- Alex Mremi: Conceptualization, Methodology, Supervision, Writing—Review & Editing.

## Acknowledgement

Our sincere appreciation should go to all the participants who generously shared their time and experiences. Special thanks to KCRI, KCMUCo, and TAWREF for their invaluable support and collaboration throughout this study.

## Declaration of AI Assistance

We hereby declare that AI technology (ChatGPT) is partially used in some areas to improve the readability and language of this work, without modifying the core ideas or content. All research tasks, data analysis, findings, and interpretations presented in this document are entirely based on our original data and were independently conducted by us.

